# The impact of cirrhosis on the inflammatory milieu before and long-term after hepatitis C virus elimination by direct-acting antiviral therapy

**DOI:** 10.1101/2023.10.31.23297828

**Authors:** Moana Witte, Carlos Oltmanns, Jan Tauwaldt, Hagen Schmaus, Jasmin Mischke, Gordon Grabert, Mara Bretthauer, Katja Deterding, Benjamin Maasoumy, Heiner Wedemeyer, Tim Kacprowski, Anke R.M. Kraft, Markus Cornberg

## Abstract

**Background and Aims:** Chronic hepatitis C virus (HCV) infection can lead to cirrhosis, development of hepatocellular carcinoma (HCC) and several extrahepatic manifestations. A sustained virological response (SVR) is achieved with direct-acting antivirals (DAA) in over 95% of the patients, but sequelae do not improve in all patients, suggesting permanent biological alterations induced by HCV infection. Therefore, we investigated the influence of chronic HCV infection, viral elimination and cirrhosis on inflammatory immune mediators.

**Approach and Results:** In 102 chronic HCV patients, 46 with and 56 without cirrhosis, 92 soluble immune mediators (SIM) were measured in plasma samples at therapy start, end of treatment and long-term follow-up (median 96 weeks). 39 HBsAg positive persons with HBeAg negative infection served as controls.

At baseline, 42 SIM were altered in chronic HCV patients (adj.p <0.05). Notably, patients with cirrhosis displayed a higher frequency and severity of alterations. At long-term follow-up, the SIM profile of the non-cirrhotic patients recovered to the level of the control group, while 41 SIM remained altered in cirrhotic patients. 33 of these SIM correlated with elastography, among them SIM linked to carcinogenesis as e.g. HGF, IL8 and IL6 (KEGG Pathways hsa05202, hsa05200).

**Conclusions:** HCV-related changes in the inflammatory milieu can persist even after HCV elimination, specifically in cirrhotic patients. These changes are closely associated with liver damage and carcinogenesis. Our findings underscore the need for HCV elimination before extensive liver injury occurs and suggest further investigation of the relationship between persistent inflammatory milieu changes and long-term sequelae after HCV elimination.

**Graphical Abstract:** 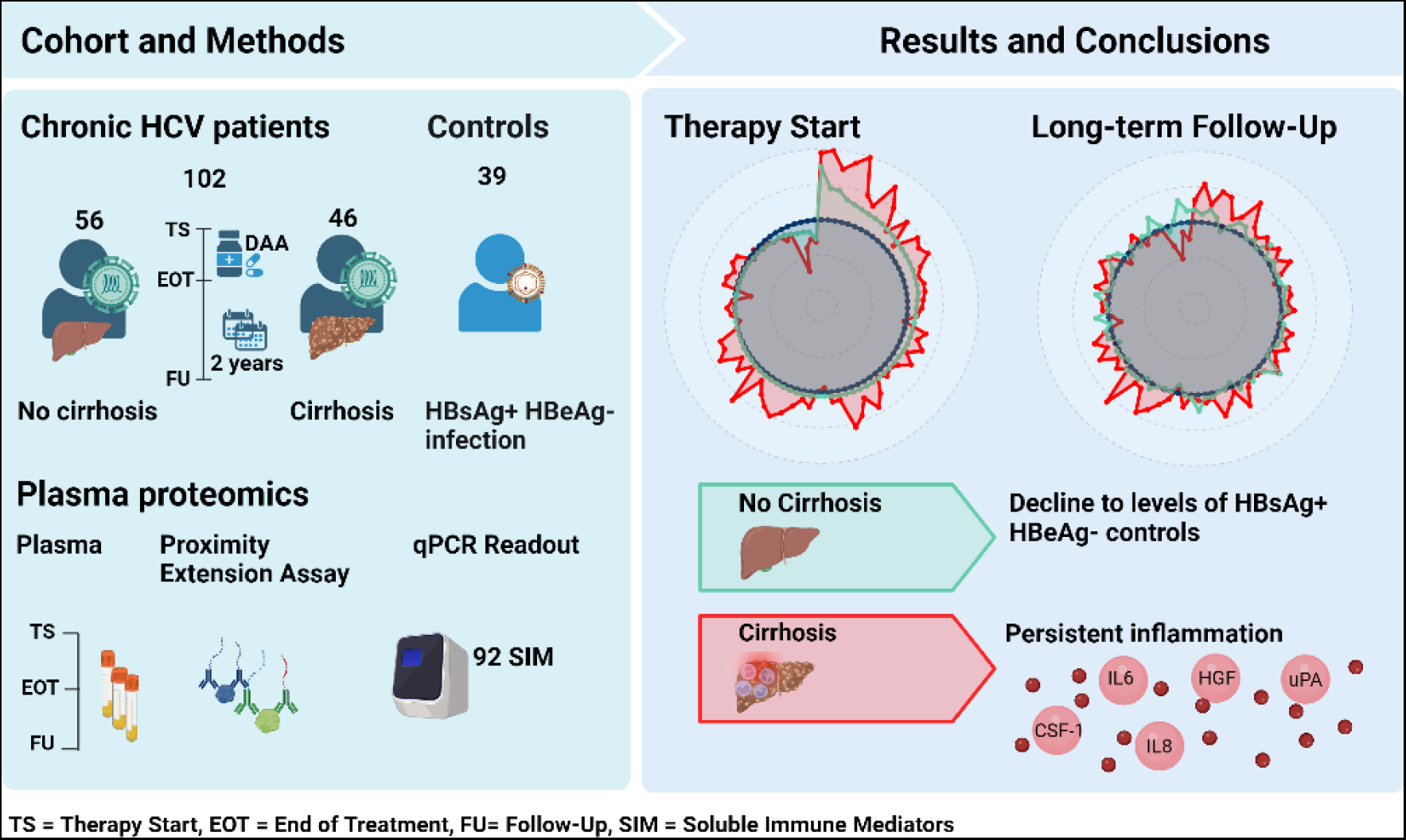

## Introduction

Hepatitis C virus (HCV) infection is one of the major causes of hepatitis and takes a chronic course in around 70% of all infected patients. In 2022, an estimated number of 58 million people were chronically infected with HCV worldwide ^1^. In addition, chronic HCV infection can lead to liver cirrhosis and hepatocellular carcinoma (HCC) ^2^ ^3^ and resulted in 290.000 deaths in 2019, being the leading cause of cirrhosis related deaths ^1^ ^4^. Besides liver-associated sequelae, chronic HCV infection also leads to several extrahepatic manifestations, e.g. lymphoma, cryoglobulinemic vasculitis, cardiovascular diseases, type 2 diabetes mellitus and fatigue in up to two thirds of patients ^5^.

Currently, chronic hepatitis C can be treated efficiently, and over 95% of patients treated with direct-acting antivirals (DAA) achieve sustained virologic response (SVR). While patients achieving SVR experience improvements in extrahepatic manifestations and a reduced risk of cirrhosis and HCC, subsequent complications and the risk of HCC are not entirely eliminated in all patients ^3^ ^6^. Especially in patients with cirrhosis and other risk factors, the risk of HCC remains high ^7^ ^8^. One possible reason for this could be biological imprints of HCV infection that are still present after SVR ^9^ ^10^. As such, HCV infection has been shown to induce epigenetic changes in the liver that persist after SVR and have been associated with the development of HCC ^11^.

Imprinting by HCV can also occur on different levels of the immune system ^9^. For example, HCV induces interferon-stimulated genes ^12^ and a characteristic inflammatory milieu that may not be fully reversible after viral elimination ^9^. Soluble inflammatory markers have been shown to remain altered for 12-24 weeks after successful DAA therapy in patients with acute ^13^ and chronic HCV infection ^14^. However, few long-term data are available on whether the changes may be reversible over the longer observation period ^13^ ^15^. As most previous studies included a heterogeneous population of patients with and without advanced liver fibrosis and cirrhosis ^14^ or did not include controls without HCV infection ^15^, we aimed to decipher the role of cirrhosis on the soluble inflammatory milieu during and after HCV infection.

Thus, we used an extensively characterized cohort of 102 chronic HCV patients with and without cirrhosis to measure 92 soluble immune mediators (SIM) at therapy start, end of treatment and long-term follow-up of 96 weeks and compared the results with 39 well-characterized healthy HBsAg positive and HBeAg negative persons with normal ALT level.

## Methods

### Study population and design

Out of 799 chronic HCV patients that were treated with DAA therapy at Hannover Medical School between January 2014 and November 2019, we selected a well-characterized cohort of 102 chronic HCV patients achieving SVR for this study. All patients were part of a prospective biobank registry and peripheral blood was collected at different time points, processed and stored after standard operating procedures. All patients gave their written informed consent for this study (Supplementary Figure 1).

We divided our patients in two groups based on non-invasive liver elastography ^16^. 46 chronic HCV patients had elastography values of above 14 kPa and were therefore included in the cirrhosis cohort (Cohort A), whereas the 56 patients that had elastography values of below 14 kPa were included in the non-cirrhosis cohort (Cohort B). For all patients, plasma samples were available at therapy start, end of treatment and long-term follow-up (median 96 weeks, Supplementary Table 1). Plasma samples from 39 well-characterized healthy HBsAg positive persons with HBeAg negative infection (all with normal ALT values) served as controls (Table 1). All controls are under long-standing follow-up in the outpatient clinic and have been evaluated for coinfections and comorbidity.

**Table 1.** Clinical characteristics at baseline of all analyzed patients, including chronic HCV patients with cirrhosis, chronic HCV patients without cirrhosis and HBsAg positive controls. Shown are mean values and standard deviations for each continuous parameter and numbers and percentages for dichotomous parameters.

| Characteristics | Cohort A<br>(cirrhosis) | Cohort B<br>(no cirrhosis) | Controls<br>(HBsAg positive) | Reference |
| --- | --- | --- | --- | --- |
| n | 46 | 56 | 39 |  |
| Age<br>(median, min, max) | 58 (± 9) | 58 (± 12) | 51 (± 10) |  |
| Sex |  |  |  |  |
| Female | 12 (26.1%) | 29 (51.8%) | 21 (53.8%) |  |
| Male | 34 (73.9%) | 27 (48.2%) | 18 (46.2%) |  |
| BMI (kg/m<sup>2</sup>) | 27.5 (±3.7) | 26.6 (± 5.2) | 26.6 (±4.2) | 18.5-24.9 |
| Elastography (kPa) | 34.6 (± 17.4) | 7.6 (± 2.5) | 5.1 (± 1.2) | 2-7 |
| Leukocytes<br>(Tsd/μl) | 5.5 (± 1.9) | 6.9 (± 1.8) | 6.4 (± 1.8) | 3.9-10.2 |
| Hemoglobin (g/dl) | 13.9 (± 1.6) | 14.8 (± 1.4) | 14.0 (± 1.2) | 13.5-17.2 (m)<br>12-15.6 (f) |
| Platelets (Tsd/μl) | 121 (± 54) | 209 (± 62) | 213 (± 47) | 160-370 |
| Neutrophils<br>(Tsd/μl) | 3.1 (± 1.1) | 4.0 (± 1.4) | 3.8 (± 1.3) | 1.5-7.7 |
| INR | 1.3 (± 0.3) | 1.0 (± 0.2) | 0.98 (± 0.4) | 0.90-1.25 |
| AST (U/L) | 119.2 (± 75) | 53.9 (± 27.7) | 25.7 (± 6.4) | 0-35 (m)<br>0-31 (f) |
| ALT (U/L) | 124 (± 93) | 73.2 (± 47.3) | 26.2 (± 15.5) | 0-45 (m)<br>0-34 (f) |
| HBV-DNA (IU/ml) |  |  | 401.3 (± 551.51) | 0 |
| HCV-RNA (IU/ml) | 1,318,155 (± 1,425,057) | 2,634,406 (± 2,972,906) |  | 0 |

All study protocols conformed to the Declarations of Helsinki and Istanbul and were reviewed and approved by the ethics committee of Hannover Medical School (No. 10873_BO_K_2023, No. 9474_BO_K_2020).

### Sample preparation

After blood draw was performed at the outpatient clinic of Hannover Medical School, all samples were processed according to standard operation procedures. EDTA whole blood was centrifuged and plasma aliquots stored at a minimum of -20 °C.

### SIM measurements

For SIM measurement, 50 µl defrosted EDTA plasma were randomized on 96 well plates and stored at -80 °C before shipping to Olink Proteomics, Upsala, Sweden. All analyses were conducted with the Target 96 Inflammation Panel including 92 SIM.

Measurements were performed in two batches. We accounted for a possible batch effect by re-analyzing 16 samples from the first batch again in the second batch (referred to as bridging samples) using the OlinkAnalyze R package ^17^. The proximity extension assay and quantitative PCR readout were performed at Olink Proteomics, Upsala, Sweden.

All data were delivered in Olink Proteomics’ arbitrary unit on a log2 scale as normalized protein expression (NPX) values. We excluded CDCP1 since the assay failed on one plate and 12 SIM that were below the limit of detection in more than 75% of all samples. A total number of 79 SIM were included in our final analysis.

The datasets generated and analyzed in the current study are available in the Synapse repository (https://www.synapse.org/#!Synapse:syn52525902).

### Statistical analysis

We performed the data analysis using R (version 4.2.2) and R Studio (version 2022.12.0). All data were normalized via bridging normalization using the OlinkAnalyze R package. Welch two sample t-tests and paired Welch two sample t-tests were used when parametrical tests were applicable. If parametrical tests were not applicable, Mann Whitney U-test and Wilcoxon signed-rank test were used. We conducted all tests using the OlinkAnalyze R package.

All p-values were adjusted for multiple testing using the Benjamini-Hochberg procedure controlling the false discovery rate (FDR) at 5%. The estimated difference in mean was determined using the OlinkAnalyze package and correlations were calculated with the rstatix package using Pearson correlation. We used multiple R packages for data visualization (Supplementary Table 2). The graphical abstract was created using Biorender.com. Network analysis was performed using StringDB (version 11.5).

## Results

### Cohort characteristics

All chronic HCV patients (n=102) achieved SVR and no HCV-RNA was detectable at long-term follow-up. Cohort A (patients with liver cirrhosis) consisted of a majority of male patients (34/46 males) with a mean age of 58 years (Table 1). As expected, liver enzymes were increased at baseline (mean AST = 119 U/L, mean ALT = 124 U/L) and a moderate thrombocytopenia (platelets = 121 Tsd/µL) was apparent. Cohort B (patients without liver cirrhosis) consisted of 56 patients (27 males) with a mean age of 58 years. Liver enzymes at baseline were increased but overall lower than in cohort A (mean AST = 53.9 U/L, mean ALT = 73.2 U/L) and platelets were not decreased (mean 209 Tsd/µL).

Thirty-nine HBsAg positive persons with HBeAg negative infection (18 males) with a mean age of 51 years served as controls. All HBsAg positive persons had low levels of HBV-DNA (<2,000 IU/ml) and no signs of liver cirrhosis or other severe liver diseases (mean AST = 25.7 U/L, mean ALT = 26.1 U/L, mean elastography = 5.1 kPa) (Table 1).

### Chronic HCV alters the SIM milieu

At therapy start, we detected a total amount of 42 altered SIM in chronic HCV patients, among others the chemokines IL8, CXCL10, CCL19 and CCL20, the growth factors HGF and SCF and several surface markers as IL-18R and PD-L1 (Figure 1, adj. p-value <0.05). Most SIM were upregulated (40/42) in comparison to the HBsAg positive controls and only TWEAK and SCF were downregulated at baseline. CXCL10, CXCL11, CCL19, CCL20 and IL8 showed highest estimated differences.

**Figure 1.**
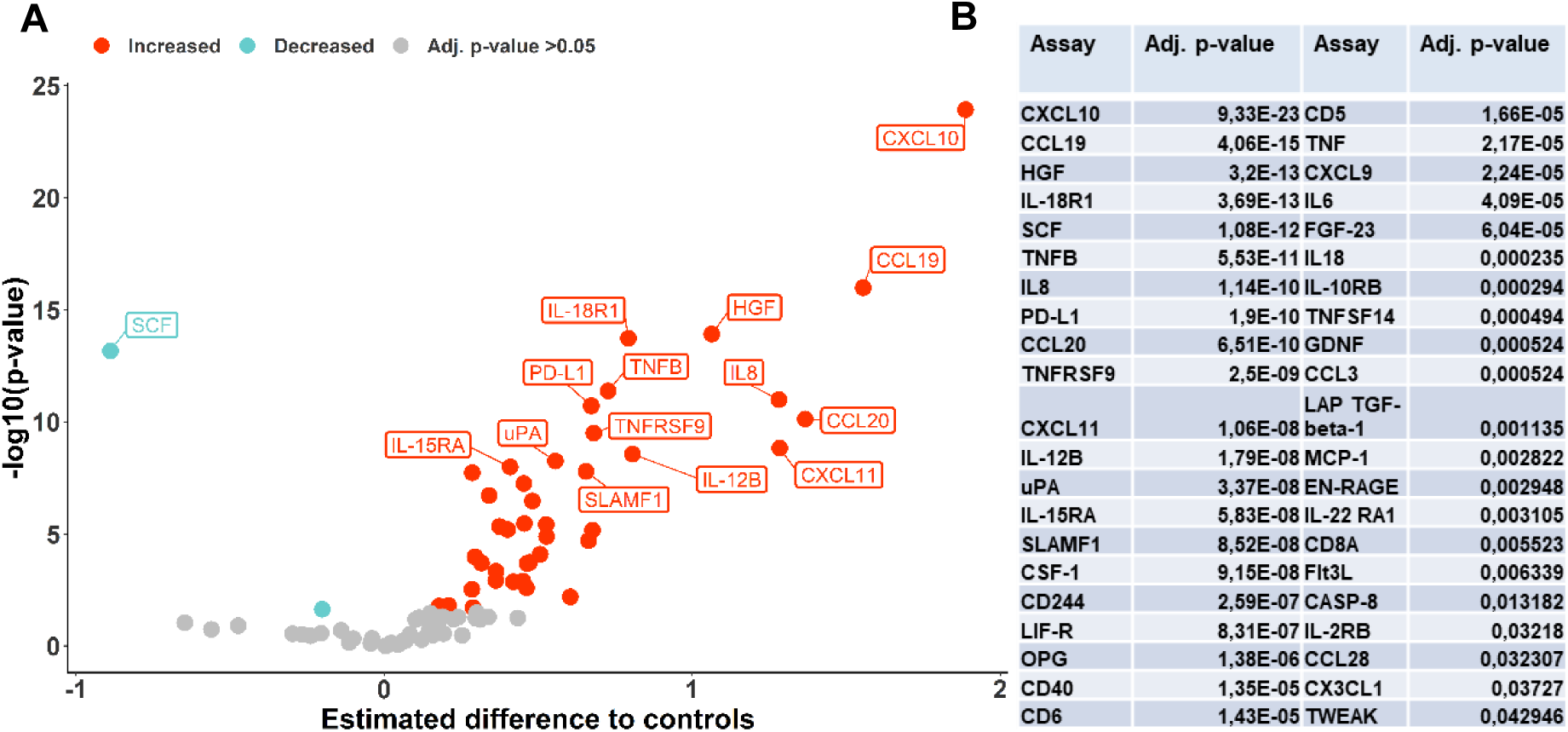
SIM expression of chronic HCV patients at baseline compared to HBsAg positive controls. (A) Significantly increased SIM are labeled in red, significantly decreased SIM in blue (adj. p-value <0.05). Non-significant SIM are represented as grey dots (adj. p-value ≥0.05). Estimated differences and p-values were calculated using Welch Two-Sample t-Test and p-values were adjusted using the Benjamini-Hochberg procedure. (B) Adjusted p-values of the 42 SIM changed in chronic HCV patients at baseline compared to controls. P-values were calculated using Welch Two-Sample t-test and adjusted using the Benjamini-Hochberg procedure.

### Cirrhotic and non-cirrhotic chronic HCV patients show distinct SIM profiles

We compared cirrhotic (cohort A) and non-cirrhotic (cohort B) patients at therapy start to the HBsAg positive controls and observed distinct SIM profiles for each cohort. We detected 50 SIM alterations in cohort A, but only 25 SIM alterations in cohort B (adj. p-value <0.05).

In order to further investigate the role of cirrhosis, we compared both SIM profiles and identified 26 out of 50 differential SIM expressions that were restricted to cirrhotic patients, including soluble surface markers such as CD5, CD6, CD244, LIF-R, IL-10RB, SLAMF1, the serine protease uPA, and ligands such as IL6 and GDNF (Figure 2A).

**Figure 2.**
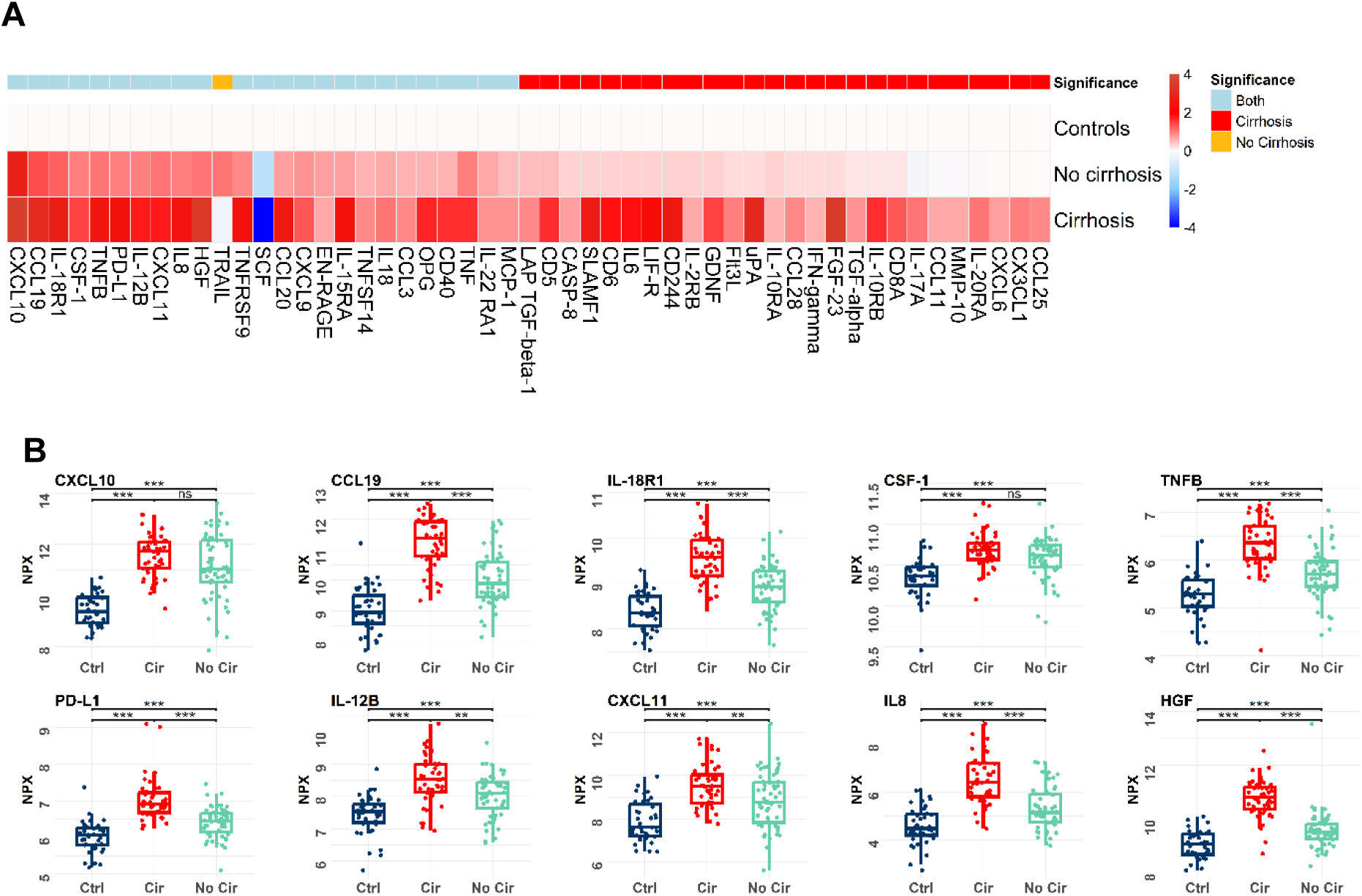
The SIM milieu of cirrhotic and non-cirrhotic patients is distinct at baseline. (A) Heatmap of significantly altered SIM in cirrhotic and non-cirrhotic patients compared to HBsAg positive controls at baseline. Shown are to controls normalized means of NPX values. Significance was determined by using Welch Two-Sample t-test and p-values were adjusted using the Benjamini-Hochberg procedure. The first row shows the significance (adj. p<0.05), SIM changed in both groups are marked in blue, SIM changed in cirrhotic patients only are red, SIM changed in non-cirrhotic patients only are yellow. (B) Boxplots of SIM altered in cirrhotic and non-cirrhotic patients at therapy start. Significance was determined by using Welch Two-Sample t-test and p-values were adjusted using false discovery rate. ***p <0.001, **p <0.01, *p <0.05, NS p >0.05.

However, 24 SIM were altered in both cohorts at baseline. Whereas 23 of these SIM, among others IL8, CXCL10, CCL19, CCL20, HGF, IL-18R1 and PD-L1, were increased, only SCF was decreased compared to the controls (Figure 2A). When we compared the expression levels of these 24 SIM in both cohorts, we detected higher NPX values for 13 of the 23 increased SIM in cirrhotic patients in comparison to non-cirrhotic patients, including IL8, CCL19, CCL20, HGF, IL-18R1, and PD-L1. Meanwhile, cirrhotic patients had lower NPX values for SCF compared to non-cirrhotic patients (Figure 2B). In consequence, cirrhotic patients (cohort A) did not only have a higher frequency of alterations at baseline, but also more severe alterations in comparison to non-cirrhotic patients (cohort B) (Figure 3).

**Figure 3.**
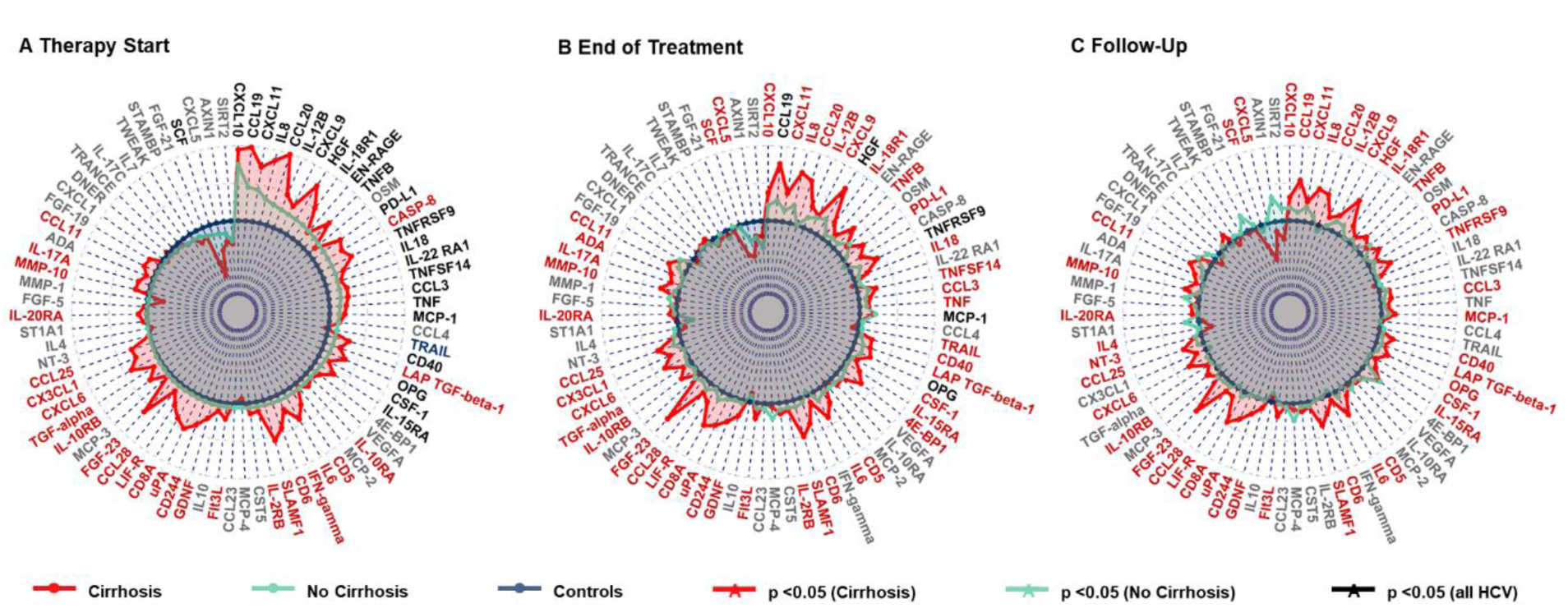
Cirrhotic and non-cirrhotic patients show distinct SIM profiles. Spidergraphs of estimated difference in means of all analyzed SIM at therapy start (A), End of Treatment (B) and Follow-Up (C). Chronic HCV patients with cirrhosis are red, chronic HCV patients without cirrhosis turquoise and HBsAg positive controls blue. Estimated differences were calculated using Welch Two-Sample t-test.

### SIM alterations in cirrhotic HCV patients persist at long-term follow-up

To investigate the influence of viral elimination on the inflammatory milieu, we further analyzed the SIM expression of our two cohorts at long-term follow up (median 96 week). In non-cirrhotic patients, no significant alterations were detectable at long-term follow-up, whereas 41 SIM alterations persisted in cirrhotic patients compared to HBsAg positive controls (Figure 3).

Among the SIM that remained altered were several chemokines as CXCL9, CXCL10, CCL11, CCL19 and CCL20, the interleukins and receptors IL6, IL8 and IL-18R1, members of the tumor necrosis factor superfamily as TNFB, CD244, CD8A, CD5 and CD6 and the growth factors HGF and SCF (Figure 3). On top of SIM that differed exclusively in cirrhotic patients, also 19/25 SIM altered in non-cirrhotic patients at baseline remained altered in cirrhotic patients at follow-up (Figure 4A).

**Figure 4.**
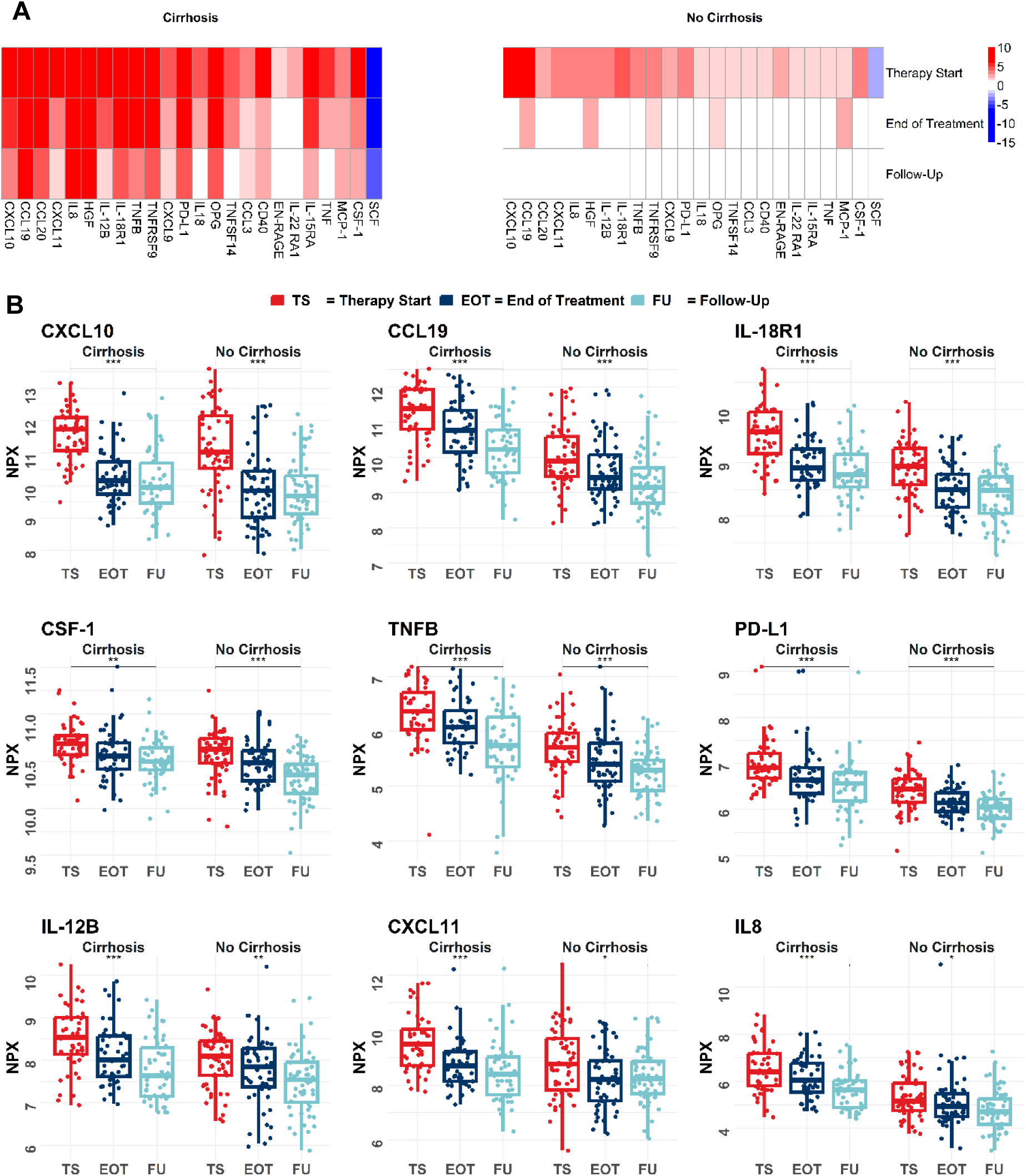
SIM alterations that persist in cirrhotic patients at follow-up. (A) Heatmap of the 24 SIM that were altered in both cohorts compared to HBsAg positive controls at baseline. Shown are significance results for all three time points compared to controls. Significance was determined by using Welch Two-Sample t-test and p-values were adjusted using the Benjamini-Hochberg procedure. (B) NPX values from cirrhotic and non-cirrhotic chronic HCV patients of 9 SIM (based on lowest p-value) that are significantly altered in cirrhotic patients at long-term follow-up. Samples were measured at therapy start (red), end of treatment (dark blue) and follow-up (light blue). Significance was determined by using paired Welch-Two Sample t-test and p-values were adjusted using the Benjamini-Hochberg procedure. ***p <0.001, **p <0.01, *p <0.05, NS p >0.05

In addition, we detected that the levels of most altered SIM in cirrhotic patients declined, but only partially and not to the levels of the controls (Figure 4B). Similar trends were also visible in the clinical data: Even though the liver enzymes AST and ALT and elastography values decreased, levels remained high in most of our cirrhotic patients (Supplementary Figure 2).

### Elastography is associated with an impaired SIM restoration

We assessed differences in SIM restoration by dividing the cohort of cirrhotic HCV patients into one group of patients that showed signs of liver fibrosis but not cirrhosis at follow-up (elastography <14 kPa, n=15) and one group of patients with persisting signs of liver cirrhosis (elastography ≥14 kPa, n=29) (Supplementary Figure 3). Both groups had similar SIM profiles at baseline, but differed at long-term follow up (Supplementary Figure 4). Whereas the inflammatory pattern persisted in the patients with high elastography at follow-up, the pattern disappeared in patients below the cut-off at follow-up (Figure 5A).

**Figure 5.**
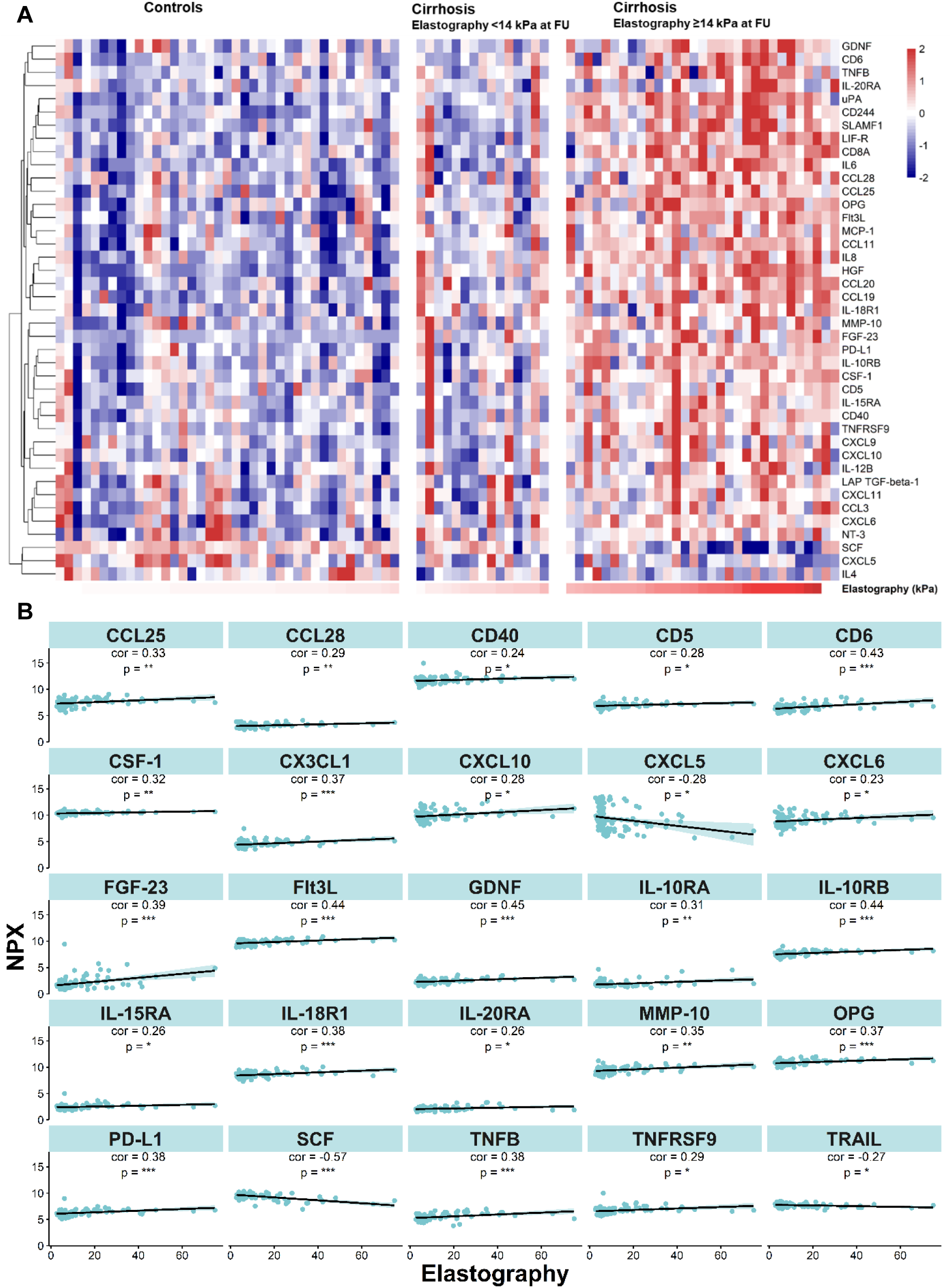
Persistent alterations are associated with elastography. (A) Heatmap of SIM alterations at follow-up in HBsAg positive controls and patients defined as cirrhotic before DAA therapy (initiation of therapy). Patients with cirrhosis at baseline were categorized into two groups based on their elastography values at follow-up (n=15 <14 kPa; n=29 ≥14 kPa). Shown are row-wise z-score normalized NPX values. (B) Top 12 correlations between NPX levels and elastography values in all HCV patients at follow-up. Correlations were calculated using Pearson correlation, p-values were adjusted using the Benjamini-Hochberg procedure. ***p <0.001, **p <0.01, *p <0.05, NS p >0.05

We detected correlations between most of the SIM (33 of 41) that remained altered in all cirrhotic patients at follow-up and the corresponding elastography values. Correlations of increased SIM were positive, while decreased SIM (e.g. SCF) were negatively correlated with elastography (Figure 5B, Supplementary Figure 5).

We aimed to decipher the interactions among the 33 SIM that remained altered in cirrhotic patients and correlated with elastography and therefore performed a network analysis using the STRING database. 26 of 33 SIM were connected on high confidence level (>0.70) with IL6 in a central position (18 edges to other SIM) (Figure 6).

**Figure 6.**
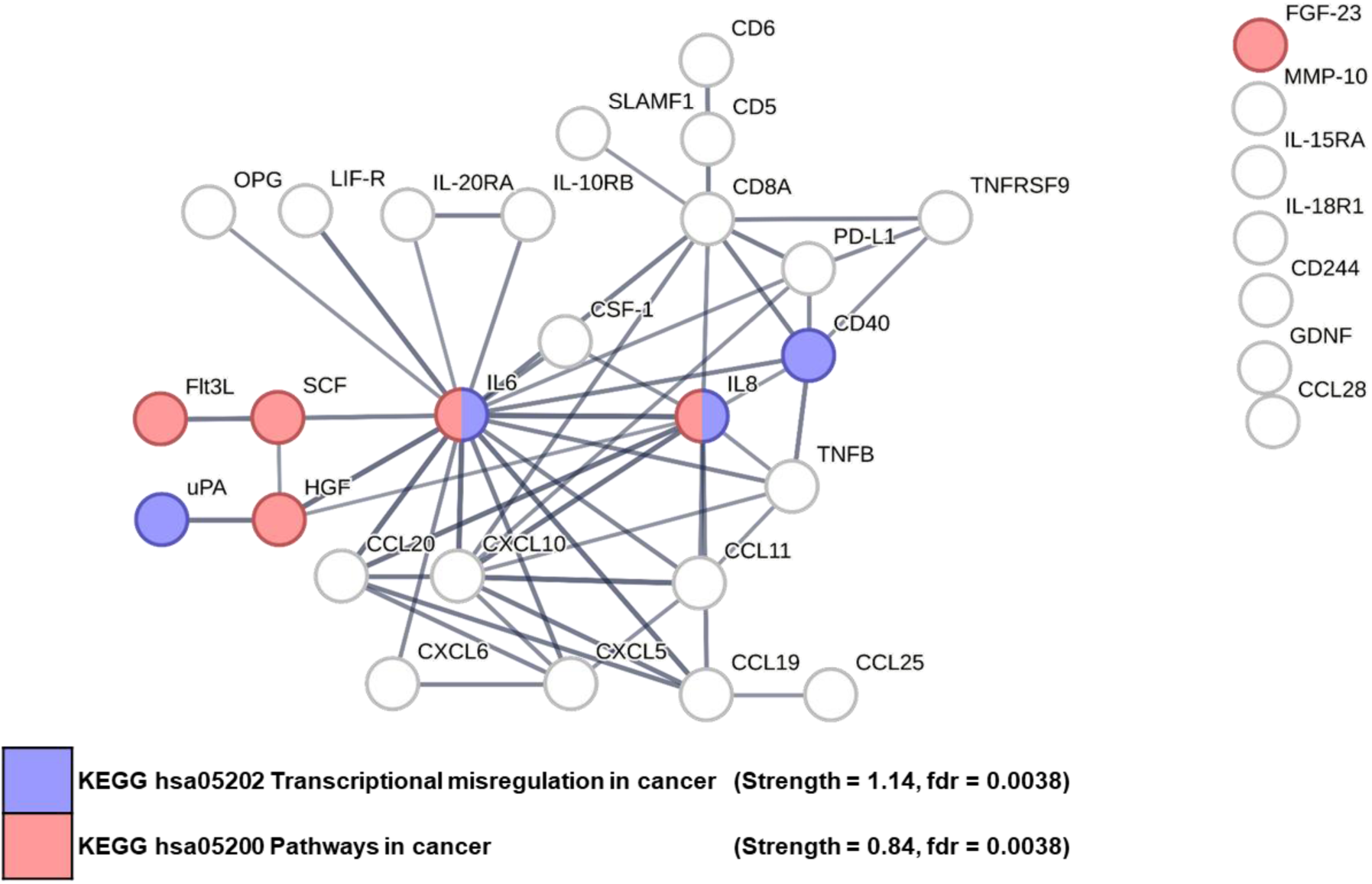
Network interaction analysis of persistently altered SIM correlating with elastography. Network was created via StringDB, edges are shown based on confidence (>0.7). The strength describes the enrichment effect based on log10 (observed/expected).

### Persistently altered SIM in cirrhotic patients associated with carcinogenesis

Based on our network analysis, we performed a functional enrichment analysis focusing on KEGG pathways. Enriched pathways were associated with inflammation and immune responses, including IL17 signalling, TNF signalling and the Toll-like receptor pathway. Major pathways were linked to NF-kappa B and PI3K-Akt signalling. Interestingly, most of the pathways included IL6 and IL8. Moreover, we detected that 7 of the significantly altered SIM were linked to pathways associated with cancer or transcriptional misregulation in cancer, among them IL6, IL8, HGF, CD40, SCF, Flt3L and uPA (Figure 7). The NPX levels of uPA, IL6, IL8, HGF, CD40 and Flt3L remained high in cirrhotic patients at long-term follow-up (Supplementary Figure 6).

**Figure 7.**
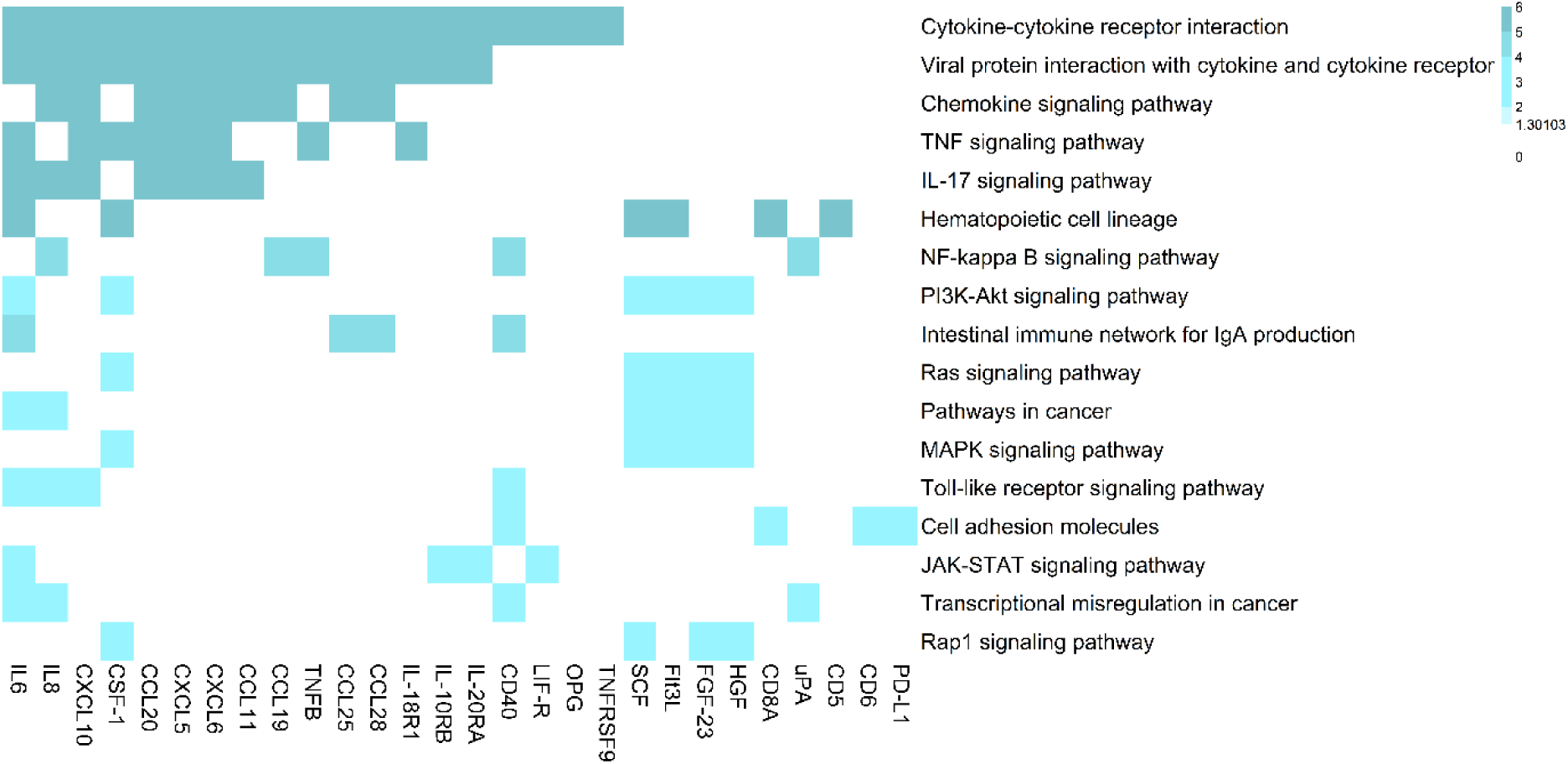
KEGG pathway enrichment analysis of altered SIM correlating with elastography. Enrichment significance (–log10 of Benjamini-Hochberg adjusted p-values) is plotted.

## Discussion

Chronic HCV infection leads to substantial alterations of the inflammatory milieu, and previous studies raised concerns about persistent changes after viral elimination through DAA treatment^18^. Our study shows a distinctive pattern in which non-cirrhotic HCV patients experienced restored SIM profiles during long-term follow-up, whereas cirrhotic patients retained persistent alterations. These results provide new insights into the lasting effects of HCV infection as well as the dynamics of immune regeneration after HCV elimination and underscore the profound impact of cirrhosis in this context. Even though HCV elimination resulted in an overall decline of inflammatory milieu alterations, only non-cirrhotic patients showed a restoration to the levels observed in controls, while 41 SIM remained altered in cirrhotic patients. Of note, most of the persistently altered SIM in patients with cirrhosis correlated with liver elastography, suggesting that the extent of liver injury is related to the degree of changes in the inflammatory milieu, which has been described previously on a smaller scale ^19^. For example, Shah et al. were able to show that IL6 is associated with the severity of liver fibrosis in patients with chronic HCV infection ^20^.

Importantly, elevated levels of IL6 but also IL8, CXCL10, uPA, and HGF play a central role in our network analysis and are recognized as pivotal factors in liver regeneration, fibrosis, and cirrhosis ^21^. *Filliol et al*. recently highlighted the central role of hepatic stellate cells in this process, showing opposing effects of HCC promotion and protection in different subpopulations ^22^. The exact underlying mechanism remains elusive, as it is likely that both pro- and anti-carcinogenic effects are active at the same time. Several studies have shown that in liver injury, the secretion of IL6 by Kupffer cells is increased. In a second step, IL6 induces the secretion of hepatocyte growth factor (HGF) by the stellate cells of the liver and is later activated by uPA, leading to proliferation of hepatocytes ^23^ ^24^. In addition, liver injury leads to the activation of hepatic stellate cells and recruitment of further inflammatory cells through secretion of cytokines as IL8 and CXCL10 that also remained altered in the cirrhotic patients^25^. IL6, IL8, HGF, CD40, SCF, Flt3L and uPA were all associated with carcinogenesis according to the KEGG analysis and should be considered in future studies as potential biomarker candidates in chronic HCV patients after cure^24 26^. Previous studies showed increased plasma or serum levels in patients with HCC after DAA therapy, but validation is lacking and no specific biomarker emerged from those studies ^27^ ^28^.

The strengths of our study include a sizeable prospective biobank cohort comprising 102 chronic HCV patients, comprehensive clinical characterization, a long follow-up period, and the assessment of 79 SIMs. However, our study also has limitations resulting from the retrospective design of the analysis and the inclusion of a control group with low replicative chronic HBV infection. Notably, all controls had low HBV-DNA, normal liver enzyme levels, no signs of liver fibrosis and underwent extensive screening for other viral infections, comorbidities, and cancer rendering them a well-studied control group. However, the potential effects of low-replicative HBV infection cannot be entirely dismissed, despite previous evidence suggesting that HBV does not directly stimulate interferon-stimulated genes (ISGs) or other innate immune responses ^29^ . Thus, our study does not provide definitive proof that the inflammatory milieu is fully restored in non-cirrhotic patients. Nevertheless, our data are consistent with recent studies focusing on non-cirrhotic chronic HCV patients that demonstrated decreased ISG expression after viral elimination ^30^. On the other hand, other previous studies suggested that cytokine alterations may still persist in all chronic HCV patients^31^. However, in addition to fibrosis and cirrhosis, other factors may also influence the regeneration of the inflammatory milieu after HCV elimination. For example, a recently published study has observed different SIM changes in chronic HCV patients after recovery depending on the degree of steatosis ^15^.

In summary, the presented data underscore the importance of early DAA therapy before the onset of cirrhosis to prevent sequelae, including persistent changes in the inflammatory milieu. Future studies should focus on the complex immune mechanisms involved in liver regeneration and carcinogenesis to identify novel therapeutic targets for patients who do not recover from cirrhosis after viral elimination, as well as predictive biomarkers for the development of HCC.

## Data Availability

The datasets generated and analyzed in the current study are available in the Synapse repository.

https://www.synapse.org/#!Synapse:syn52525902

## Acknowledgments

We thank Helena Lickei for her assistance with blood sample processing. We thank the study nurses (Neslihan Devici, Carola Mix, Janet Cornberg, Jennifer Witt, Julia Schneider) and the physicians (Christopher Dietz, Kerstin Port, Tammo Tergast) of the Hepatitis Outpatient Clinic of the Department of Gastroenterology, Hepatology, Infectious Diseases and Endocrinology of Hannover Medical School for the care of the patients in the patient registry. We thank all patients for participating in our research study and for donating blood. We thank EASL for the opportunity to present preliminary data at the EASL congress 2023 in Vienna.

## Notes

**Financial support and sponsorship:** This work was supported by the German Federal Ministry of Education and Research (BMBF) within the framework of the CompLS research and funding concept (grants 031L0294C, 031L0294A). The project was supported by infrastructure of the German Center for Infection Research (DZIF; TTU 05.708_00, TTU-IICH-07-808) and the Cluster of Excellence Resolving Infection Susceptibility (RESIST; EXC 2155). This project was part of project A5 in the Collaborative Research Center 900 - Microbial Persistence and its Control. This study and MW were supported by the Else Kröner-Fresenius-Stiftung (Promotionsprogramm DigiStrucMed 2020_EKPK.20). CO, JT were supported by by the Else Kröner-Fresenius-Stiftung (Promotionsprogramm KlinStrucMed).

**Conflicts of interest:**MW, CO, JT, HS, JM, GG, MB, AK and TK have nothing to disclose. KD reports research grants and personal fees from AbbVie, Alnylam and Gilead, outside the submitted work. BM received speaker and/or consulting fees from Abbott Molecular, Astellas, Intercept, Falk, AbbVie, Luvos, Norgine, Gore, Gilead, Fujirebio, Merck (MSD), and Roche. He also received research support from Abbott Molecular, Altona Diagnostics, EWIMED, Fujirebio and Roche, outside the submitted work. HW reports grants/research support and personal fees from Abbvie, Biotest AG and Gilead. He received personal fees from Aligos Therapeutics, Altimmune, Astra Zeneca, Bristol-Myers- Squibb, BTG Pharmaceuticals, Dicerna Pharmaceuticals, Enanta Pharmaceuticals, Dr. Falk Pharma, Falk Foundation, Intercept Pharmaceuticals, Janssen, Merck KGaA, MSD Sharp & Dohme GmbH, MYR GmbH, Norgine, Novartis, Pfizer Pharma GmbH, Roche and Vir Biotechnology, outside the submitted work. MC reports honoraria for lectures and consulting from AbbVie Deutschland GmbH & Co. KG, AiCuris AG, Falk Foundation e.V., Gilead Sciences GmbH, GSK Service Unlimited, MSD Sharp & Dohme GmbH, Novartis AG, Roche AG, Spring Bank Pharmaceuticals, Swedish Orphan Biovitrum AB (SOBI), outside the submitted work.

### Competing Interest Statement

MW, CO, JT, HS, JM, GG, MB, AK and TK have nothing to disclose.
KD reports research grants and personal fees from AbbVie, Alnylam and Gilead, outside the submitted work.
BM received speaker and/or consulting fees from Abbott Molecular, Astellas, Intercept, Falk, AbbVie, Luvos, Norgine, Gore, Gilead, Fujirebio, Merck (MSD), and Roche. He also received research support from Abbott Molecular, Altona Diagnostics, EWIMED, Fujirebio and Roche, outside the submitted work.
HW reports grants/research support and personal fees from Abbvie, Biotest AG and Gilead. He received personal fees from Aligos Therapeutics, Altimmune, Astra Zeneca, Bristol-Myers-Squibb, BTG Pharmaceuticals, Dicerna Pharmaceuticals, Enanta Pharmaceuticals, Dr. Falk Pharma, Falk Foundation, Intercept Pharmaceuticals, Janssen, Merck KGaA, MSD Sharp & Dohme GmbH, MYR GmbH, Norgine, Novartis, Pfizer Pharma GmbH, Roche and Vir Biotechnology, outside the submitted work.
MC reports honoraria for lectures and consulting from AbbVie Deutschland GmbH & Co. KG, AiCuris AG, Falk Foundation e.V., Gilead Sciences GmbH, GSK Service Unlimited, MSD Sharp & Dohme GmbH, Novartis AG, Roche AG, Spring Bank Pharmaceuticals, Swedish Orphan Biovitrum AB (SOBI), outside the submitted work.

### Funding Statement

This work was supported by the German Federal Ministry of Education and Research (BMBF) within the framework of the CompLS research and funding concept (grants 031L0294C, 031L0294A). The project was supported by infrastructure of the German Center for Infection Research (DZIF; TTU 05.708_00, TTU-IICH-07-808) and the Cluster of Excellence Resolving Infection Susceptibility (RESIST; EXC 2155). This project was part of project A5 in the Collaborative Research Center 900 - Microbial Persistence and its Control. This study and MW were supported by the Else Kroener-Fresenius-Stiftung (Promotionsprogramm DigiStrucMed 2020_EKPK.20). CO, JT were supported by by the Else Kroener-Fresenius-Stiftung (Promotionsprogramm KlinStrucMed).

