## Supplementary Material for "The impact of cirrhosis on the inflammatory milieu before and long-term after hepatitis C virus elimination by direct-acting antiviral therapy"

### Supplementary Tables

**Supplementary Table 1:** Samples of chronic HCV patients included in the analysis

| | **Cohort A (cirrhosis)** | **Cohort B (no cirrhosis)** |
|---|---|---|
| Treatment start (TS) | 46 | 56 |
| End of treatment (EOT) | 46 | 56 |
| Follow-up (FU) in weeks | 38 FU96<br>3 FU24<br>3 FU48<br>1 FU192<br>1 FUY240 | 56 FU96 |

**Supplementary Table 2:** R packages used for data analysis and visualization

| |
|---|
| **janitor_2.2.0** |
| **rstatix_0.7.2** |
| **msigdbr_7.5.1** |
| **gplots_3.1.3** |
| **scales_1.2.1** |
| **gridExtra_2.3** |
| **cowplot_1.1.1** |
| **ggradar_0.2** |
| **paletteer_1.5.0** |
| **fmsb_0.7.5** |
| **writexl_1.4.2** |
| **ggpubr_0.6.0** |
| **ggsignif_0.6.4** |
| **RColorBrewer_1.1-3** |
| **ggvenn_0.1.9** |
| **ggVennDiagram_1.2.2** |
| **readxl_1.4.2** |
| **pheatmap_1.0.12** |
| **clusterProfiler_4.6.0** |
| **OlinkAnalyze_3.3.1** |
| **lubridate_1.9.2** |
| **forcats_1.0.0** |
| **stringr_1.5.0** |
| **dplyr_1.1.0** |
| **purrr_1.0.1** |
| **readr_2.1.4** |
| **tidyr_1.3.0** |
| **tibble_3.2.0** |
| **tidyverse_2.0.0** |
| **ggrepel_0.9.3** |
| **ggplot2_3.4.1** |

### Supplementary Figures

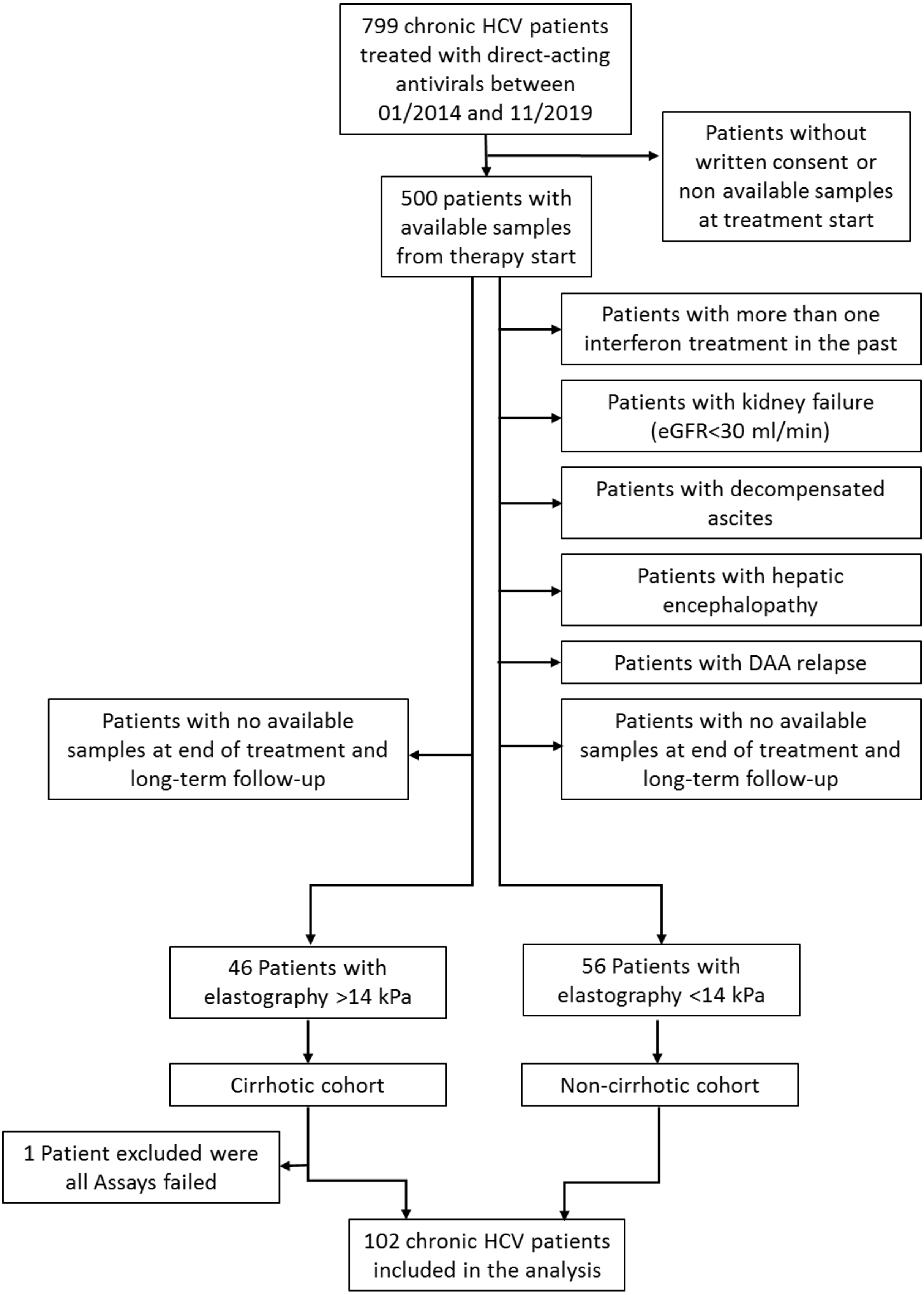


**Supplementary Figure 1**: Criteria for inclusion and exclusion of HCV patients

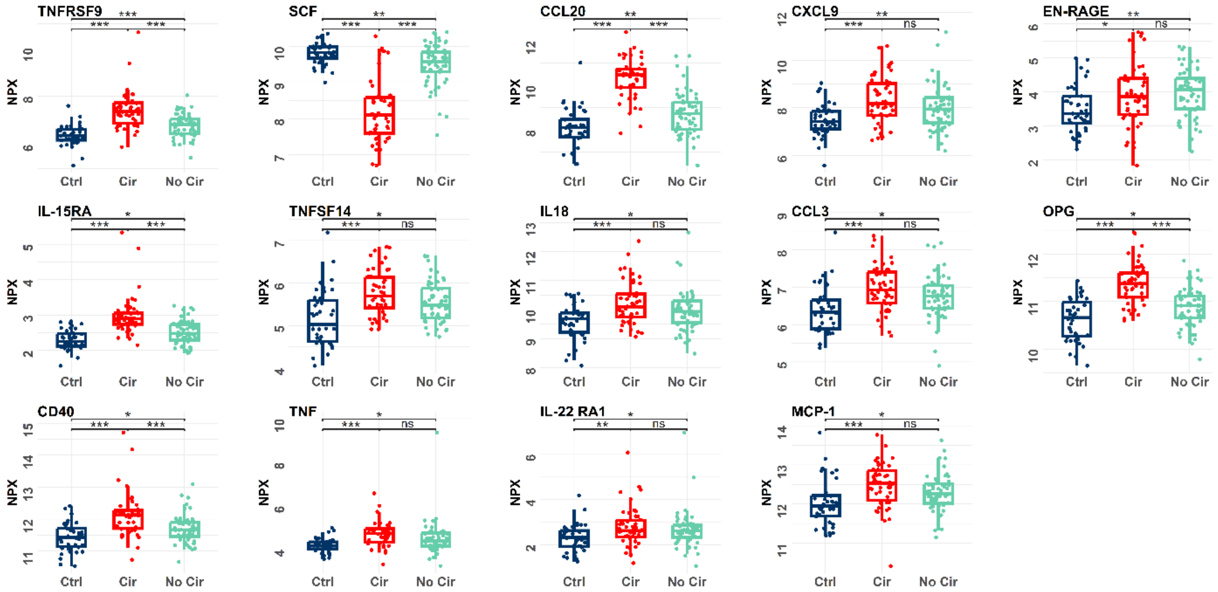


**Supplementary Figure 2:** Additional Boxplots of SIM altered in both cohorts at therapy start. Significance was determined by using Welch Two-Sample t-test and p-values were adjusted using the Benjamini-Hochberg procedure. HBsAg positive Controls (blue), Cirrhosis (red), no Cirrhosis (turquoise). ***p <0.001, **p <0.01, *p <0.05, NS p >0.05

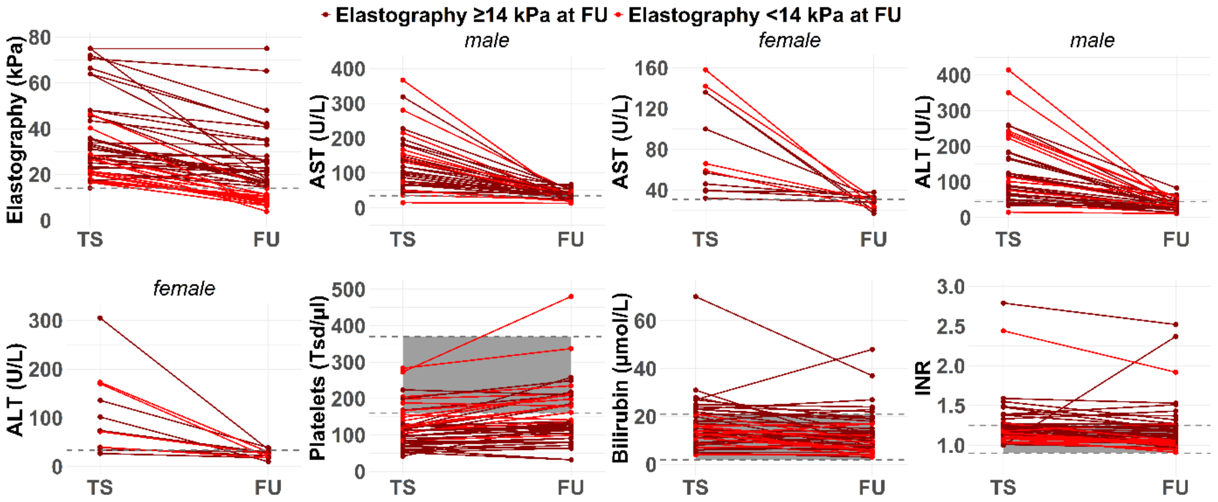


**Supplementary Figure 3:** Selected clinical parameter in cirrhotic HCV patients at therapy start and follow-up. Shown are clinical parameters for patients with elastography ≥14kPa at follow up (darkred) and <14kPa at follow up (red). Reference thresholds or areas are marked in grey.

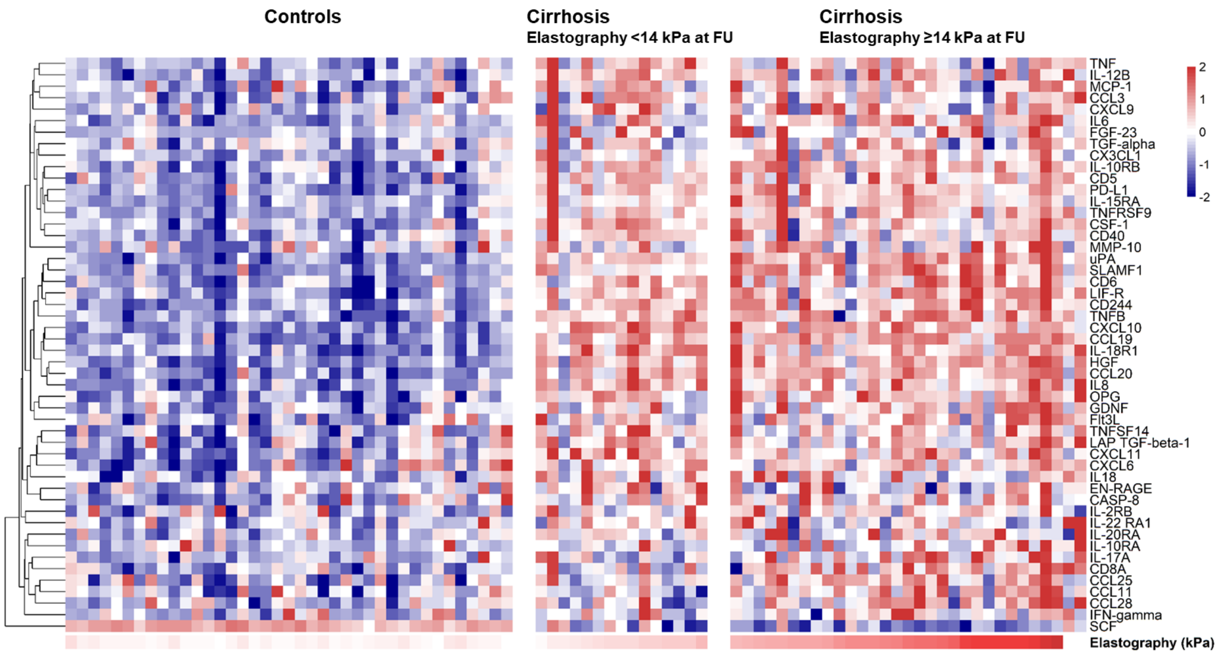


**Supplementary Figure 4:** Heatmap of SIM alterations before start of DAA therapy in HBsAg positive controls and patients defined as cirrhotic before DAA therapy (initiation of therapy). Patients with cirrhosis at baseline were categorized into two groups based on their elastography values at follow-up (n=15 ≥14 kPa; n=29 >14 kPa). Shown are row-wise z-score normalized NPX values.

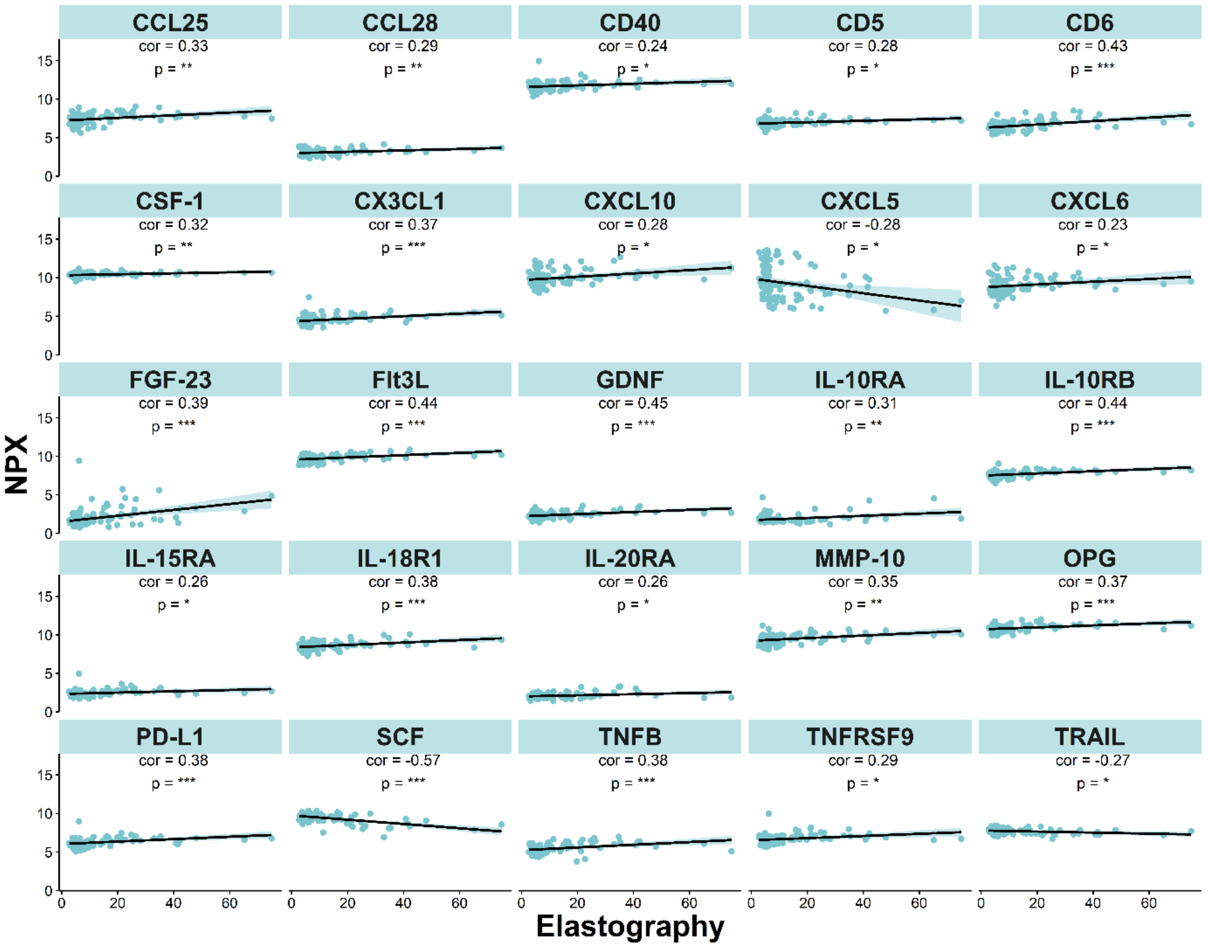


**Supplementary Figure 5:** Additional correlating SIM in all HCV patients at follow-up. Correlations were calculated using two-sided Pearson correlation, p-values were adjusted using the Benjamini-Hochberg procedure. ***p <0.001, **p <0.01, *p <0.05, NS p >0.05

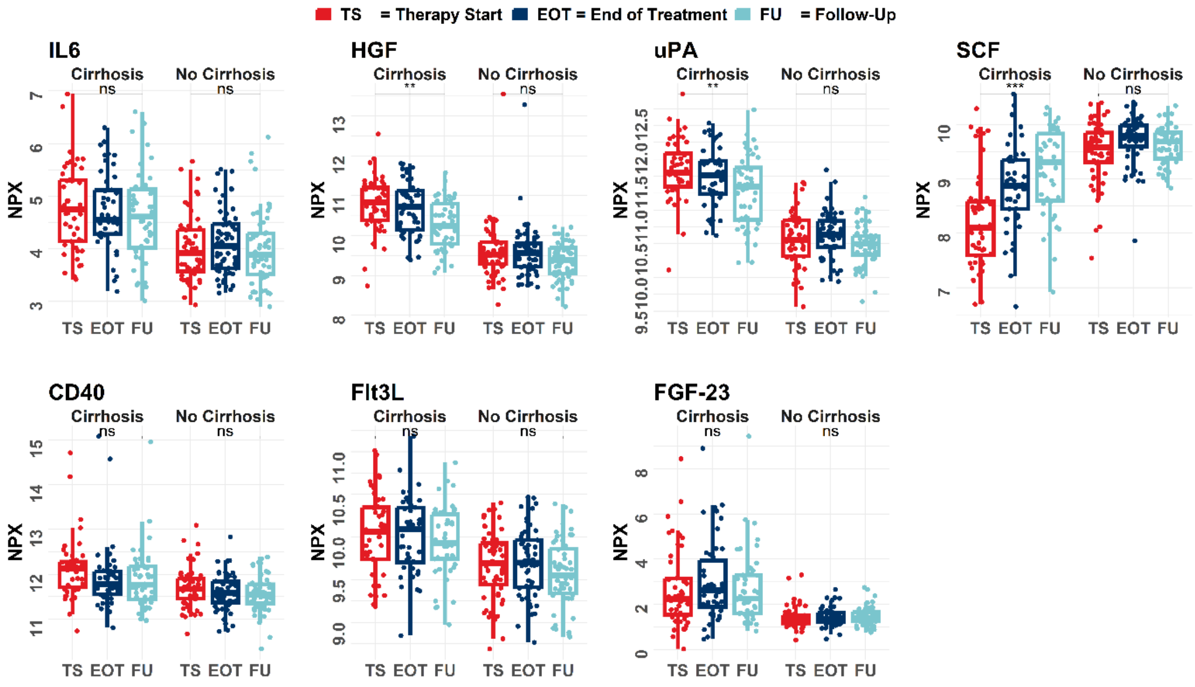


**Supplementary Figure 6:** Additional boxplots of SIM associated with carcinogenesis.

Significance was determined by using Welch Two-Sample t-test and p-values were adjusted using the Benjamini-Hochberg procedure. Shown are values of therapy start (red), end of treatment (blue), follow-up (lightblue), ***$p < 0.001$, **$p < 0.01$, *$p < 0.05$, NS $p > 0.05$
